# Cross-cohort validation and cutpoint estimation of the Janssen plasma p-tau217+ assay in predominantly cognitively normal community cohorts

**DOI:** 10.1101/2025.10.02.25331921

**Authors:** Wasiu G. Balogun, Xuemei Zeng, Gallen Triana-Baltzer, Anum Saeed, Happiness E. Aigbogun, Alexandra Gogola, Brian J. Lopresti, Victor L Villemagne, Mary Ganguli, Hartmuth Kolb, Beth E. Snitz, Oscar L. Lopez, Ann D. Cohen, Steven E. Reis, Thomas K. Karikari

## Abstract

**INTRODUCTION:** Cross-cohort validation studies for plasma p-tau217 are limited. We evaluated the Janssen plasma p-tau217+ assay and proposed a cutpoint value in three independent community-based cohorts.

**METHODS:** We included n=441 participants (age=70.3±7.3), with Aβ-PET, tau-PET, clinical and cognitive information.

**RESULTS:** The cohorts had low pre-test probability (%Aβ positivity=14.9–24.7) and were predominantly cognitively normal (>73%). Plasma p-tau217+ had high accuracy for abnormal Aβ PET (AUCs=81-86%), good correlation with Aβ-PET burden (0.336-0.397) that was highest in the cohort with the most Aβ-PET-positive participants, and the biomarker concentrations were highest in the joint Aβ-PET and tau-PET positive group. NPV was high across cohorts (≤93%) but PPV was consistently poor (<57%). Sensitivity and specificity averaged 75% and 84% respectively. A combined cohort cutpoint of 0.05pg/ml gave AUC=84.5%, NPV=94%, PPV=50%, sensitivity=75%, and specificity=84%.

**DISCUSSION:** Plasma p-tau217+ can rule out Aβ pathophysiology due to Alzheimer’s disease at the population level. Cohort-level %Aβ-PET positivity influences accuracies.

## 1 INTRODUCTION

The use of cerebrospinal fluid (CSF) and imaging techniques like positron emission tomography (PET) for the antemortem diagnosis of Alzheimer’s disease (AD), has led to significant advancements in the field.^1–3^ This has enabled the detection of AD in cognitively unimpaired individuals years before symptom onset, and has provided a foundation for interventions, particularly anti-amyloid interventions, which are known to remove Aβ plaques from the brain.^2–5^ These techniques have brought hope in the fight against AD. However, there are limitations to these approaches, including invasiveness, cost, and the need for specialized expertise.^2,3,6^ As a result, researchers have developed blood-based biomarkers as potential alternatives. These biomarkers show promise for diagnosing and predicting AD and are currently being considered for clinical use. In May 2025, the FDA cleared the Lumipulse G pTau217/β-Amyloid 1-42 Plasma Ratio for clinical use, marking it as the first blood-based biomarker for the early detection of amyloid plaques.

Several blood-based biomarkers are gaining popularity in AD research, such as glial fibrillary acidic protein (GFAP), ratio of Amyloid beta 42 to Amyloid beta 40 (Aβ42/Aβ40 ratio), Neurofilament light chain (NfL), phosphorylated tau181 (p-tau181), phosphorylated tau217 (p-tau217) and phosphorylated tau231 (p-tau231).^1,7–9^ These biomarkers are being extensively studied for their potential applications in memory clinics, primary care settings, and in clinical trials.^7,9^ Among the tau biomarkers, p-tau217 has shown the most consistent and most promising results. The plasma p-tau217 assay has been found to perform similarly to the CSF p-tau217 assay in discriminating between AD and other neurodegenerative disorders,^3,10–11^ making it a promising but less invasive alternative. This has led to its widespread acceptance, resulting in different diagnostic companies using various techniques, including mass spectrometry and immunoassay to develop assays for measuring p-tau217 in blood. Most plasma p-tau217 assays utilize immunoassays.^12,13^ The Janssen p-tau217+ assay, developed by Janssen Research and Development, uses the Single molecule immunoassay (SIMOA) technology to quantify minute amounts of p-tau217 in human blood samples. Unlike other assays, this method specifically uses the PT3 antibody which has a unique binding affinity for tau protein phosphorylated at Thr217. The binding is further enhanced when there is simultaneous phosphorylation at the neighboring sites Thr212 and Ser214, hence the name p-tau217+.^14,15^ The Janssen p-tau217+ assay is currently being marketed as a Laboratory Developed Test by the Quanterix Corporation subsidiary diagnostic company Lucent Diagnostics. Since this assay is currently being used clinically, it is necessary to widely validate it in community-dwelling older adults who are less likely to join classical research cohorts.

While there have been studies evaluating the technical performance and ability of the Janssen p-tau217+ assay to monitor and detect the pathology of AD, especially in Aβ positive participants, and comparing it to other commercially available plasma p-tau217 assays,^7,15,16^ there is a lack of studies that assess the p-tau217+ assay in community-based cohorts with particular interest in the preclinical AD phase. Therefore, this study aimed to investigate the association of the Janssen plasma p-tau217+ assay with Aβ PET and tau PET and its classification accuracies across three largely cognitively normal community cohorts. Additionally, we took advantage of the multicohort approach to generate and cross-validate preliminary cutpoints for plasma p-tau217+.

## 2 METHODS

### 2.1 Cohort studies and blood sampling

In this study, three community-based independent cohort studies representing diverse groups of participants were used. The methods have been described extensively previously.^17–21^

#### Cohort 1 (MYHAT-NI)

The Monongahela Youghiogheny Healthy Aging Team-Neuroimaging (MYHAT-NI) is a sub-cohort of the parent MYHAT study, which is a community-based sample of older adults without dementia, designed to investigate mild cognitive impairment in older adults in an economically depressed Rust Belt region in southwestern Pennsylvania.^17,18^ The parent MYHAT study recruited participants aged 65 and above, utilizing age-stratified random sampling from voter registration lists publicly available . The MYHAT-NI study recruited a subgroup of MYHAT participants who had a Clinical Dementia Rating (CDR) sum of box score of < 1.0 for neuroimaging assessments to study the distribution and functional correlations of AD pathologies. For this reason, all MYHAT-NI participants had normal or only very mildly impaired cognition at the time of enrollment. The only exclusion criterion was a contraindication to neuroimaging. Sociodemographic information and cognitive test data were collected at the parent study visit. Blood samples were collected during the baseline neuroimaging visit, which included [^11^C] Pittsburgh Compound B (PiB) positron emission tomography (PET) imaging of Aβ plaques and [^18^F]AV-1451 PET imaging of tau neurofibrillary tangles. Detailed study designs for MYHAT-NI, including subject recruitment strategies, multi-domain cognitive assessments, neuroimaging, *APOE* ε4 genotyping and data processing have been published by our group previously.^17–19^

#### Cohort 2 (CoBrA)

The Connectomics in Brain Aging and Dementia (CoBrA) study, under the auspices of the Human Connectome Project (HCP) is a community-based longitudinal study that aimed to recruit an equal number of self-identified Black/African American and non-Hispanic White participants aged 50-89 years old.^20^ The study aims to accelerate and address questions related to the structure and function of AD and other neurovascular diseases leveraging the comparative advantage of different imaging techniques, including MRI, magnetoencephalography, and PET imaging. The participants underwent neuroimaging assessment with [^11^C]PiB PET imaging of Aβ plaques. Demographic information was collected and neuropsychological assessment completed. Neuropsychological test battery assessments used include the Montreal Cognitive Assessment (MoCA), verbal fluency, a 30-item visual naming test, Trail making verbal free recall and the Rey-Osterreith Complex Figure to measure their motor functions, cognitive status, emotions and personality traits. Blood samples were collected and processed for biomarker analysis, DNA analysis and *APOE* ε4 genotyping. Further information about this cohort can be found in a previous publication.^20^

#### Cohort 3 (HeartSCORE)

The Heart Strategies Concentrating on Risk Evaluation (HeartSCORE) Study began in 2003 as a community-based cohort study of 2000 participants recruited in Allegheny County, Pennsylvania. Participants were 45 to 75 years old at baseline. The exclusion criteria include comorbid conditions that can limit life expectancy to <5 years and inability to undergo annual follow-up visits. Demographic and medical histories were collected including information on self-reported race, marital/co-habiting status, education, income, smoking, physical activity, and dietary habits. A subset of participants from the HeartSCORE study were recruited for neuroimaging and neurocognitive testing. All participants underwent a Mini-Mental State Examination. Detailed information about the study design for HeartSCORE, including data collection and processing, study recruitment, retention strategies, cognitive assessments, and neuroimaging can be found in previous publications.^21^

### 2.2 Neuroimaging

[^11^C]PiB PET and [^18^F]AV-1451 PET were used to assess brain Aβ and tau pathology respectively. The A status was based on a global [^11^C] PiB standardized uptake value ratio (SUVR) computed by volume-weighted averaging of nine composite regional outcomes (anterior cingulate, posterior cingulate, insula, superior frontal, cortex, orbitofrontal cortex, lateral temporal cortex, parietal, precuneus, and ventral striatum). Participants were classified as A+ or A-, with A+ reported as SUVR >1.346 and A-reported as SUVR <1.346. For T status, we used a MetaTemporal composite region^22^. Participants were classified as T+ and T-, with SUVR >1.18 considered as T+ while SUVR ≤1.18 considered as T-. For the neuroimaging analysis, this study utilized the same tracers, scanners, and protocol throughout the study to ensure consistent results.

### 2.3 Plasma Biomarker Assay

The plasma p-tau217+ assay measurements were conducted at Quanterix (Billerica, MA) using the Janssen assay (aka LucentAD p217).^7^ The plasma p-tau217+ measurements were performed as in-kind research collaboration with the University of Pittsburgh (PI Karikari) under material transfer agreement. However, the scientists conducting the measurements were blinded to the sample information until the measurements were completed. Plasma p-tau217+ measurements were performed on a Quanterix HD-X instrument, utilizing the monoclonal anti-human p-tau217 antibody PT3, the binding affinity of which towards p-tau217 is enhanced by additional phosphorylation at aa212, as the capture antibody. Anti-human tau antibody HT43 was used as the detector.^7^ Before starting the assay measurement, the plasma samples were thawed at room temperature and then centrifuged at 4000xg for 10 minutes to remove any particulate matter. The intra-assay and inter-assay %coefficients of variation (CVs) for the assay were 3.9% and 7.3% respectively.

### 2.4 Statistical Analysis

Plasma p-tau217+ data were independently unblinded and matched with the clinical, neuropsychological and neuroimaging datasets and statistical analysis performed at the University of Pittsburgh without any involvement of Janssen or Quanterix staff. All analyses were conducted using MATLAB (version R2021b) or R statistical software version 4.2.1 (R Foundation for Statistical Computing, Vienna, Austria; http://www.r-project.org/). For demographic characteristics, we presented the continuous variables as medians and interquartile ranges (IQR) and categorical variables as counts and percentages. Wilcoxon rank-sum tests were used to compare continuous variables, and Fisher’s exact tests were used for categorical variables between the A+ and A-groups. Additionally, Wilcoxon rank-sum tests were employed for comparing p-tau217+ levels dichotomized by A status, T status, and cognitive impairment. The Kruskal-Wallis test was employed to compare plasma p-tau217+ level among four groups stratified by A and T statuses. Spearman’s rank-based correlation measured the relationship between plasma p-tau217+ levels, and neuroimaging-assessed AD pathology burden. The Area Under the Curve (AUC) of the Receiver Operating Characteristic (ROC) curve, with and without covariates (age, sex, and *APOE* ε4 carrier status), was determined using linear regression, with log2-transformed p-tau217+ levels as the dependent variable. The predictive performance, including sensitivity, specificity, negative predictive value (NPV), and positive predictive value (PPV), was based on the Youden index. Linear regression models were also used to evaluate the significance of the association between p-tau217+ levels and demographic and genotypic risk factors.

## 3 RESULTS

### 3.1 Cohort Characteristics

We included a total of 441 participants from the MYHAT-NI (n=93), CoBrA (n=201) and HeartSCORE (n=147) cohorts.

The MYHAT-NI cohort had a median (IQR) age of 76 (72-81) years, 48 (51.6%) females, 89 (95.7%) non-Hispanic Whites (NHW), and 13 (14%) *APOE* ε4 carriers. Among them, 23 (24.7%) participants were A+, 33 (35.5%) were T+, 29 (31.2%) were N+, and 85 (91.4%) had no dementia (CDR global score of 0) (Table 1a). The A+ participants were older (p=0.025), had higher CDR global scores (i.e., more cognitive impairment; p=0.021), and a higher prevalence of *APOE* ε4 carriers (p<0.001) than the A-group. They also had a higher likelihood of being T+ (p<0.001). There were no significant differences in sex, education, race, MMSE scores, or N status (whether analyzed as a dichotomous or continuous variable) between the A+ and A-groups.

**Table 1:** Participant characteristics in the MYHAT-NI, CoBrA, and HeartSCORE cohorts according to Aβ PET positivity.

**a. Cohort: MYHAT-NI**
|  | MYHAT-NI COHORT |  |  |  |
| --- | --- | --- | --- | --- |
| | Total | Aβ PET- | Aβ PET+ | p-value |
| **N (%)** | 93 | 70 (75.3%) | 23 (24.7%) |  |
| **Age (years)** | 76.0 (72.0-81.0) | 76.0 (71.0-80.0) | 80.0 (76.0-83.8) | 0.025 |
| **Sex** |  |  |  | 0.344 |
| **Female** | 48 (51.6%) | 34 (48.6%) | 14 (60.9%) |  |
| **Male** | 45 (48.4%) | 36 (51.4%) | 9 (39.1%) |  |
| **Education (years)** | 12 (12-16) | 12 (12-16) | 13 (12-16) | 0.970 |
| **Race** |  |  |  | 1.000 |
| **NHW** | 89 (95.7%) | 67 (95.7%) | 22 (95.7%) |  |
| **B/AA** | 4 (4.3%) | 3 (4.3%) | 1 (4.3%) |  |
| **MMSE** |  |  |  | 0.434 |
| **$\geq 24$** | 91 (97.85%) | 69 (98.6%) | 22 (95.7%) |  |
| **19-23** | 2 (2.15%) | 1 (1.4%) | 1 (4.3%) |  |
| **CDR global score** |  |  |  | 0.021 |
| **CDR=0** | 85 (91.4%) | 67 (95.7%) | 18 (78.3%) |  |
| **CDR=0.5** | 8 (8.6%) | 3 (4.3%) | 5 (21.7%) |  |
| **APOE $\epsilon 4$ carrier** |  |  |  | <0.001 |
| **Yes** | 13 (14.0%) | 3 (4.3%) | 10 (43.5%) |  |
| **No** | 80 (86.0%) | 67 (95.7%) | 13 (56.5%) |  |
| **PiB SUVR** | 1.14 (1.01-1.33) | 1.10 (1.07-1.15) | 1.70 (1.48-1.89) | <0.001 |
| **Tau PET** |  |  |  | <0.001 |
| **Negative** | 60 (64.5) | 52 (74.3%) | 8 (34.8%) |  |
| **Positive** | 33 (35.5) | 18 (25.7%) | 15 (65.2%) |  |
| **Tau PET SUVR** | 1.16 (1.11-1.21) | 1.14 (1.09-1.18) | 1.21 (1.17-1.26) | <0.001 |

**b. Cohort: CoBrA**
|  | CoBrA COHORT |  |  |  |
| --- | --- | --- | --- | --- |
| | Total | Aβ PET- | Aβ PET+ | p-value |
| **N (%)** | 201 | 171 (85.1%) | 30 (14.9%) |  |
| **Age (years)** | 62.0 (57.0-71.0) | 60.0 (56.0-68.0) | 71.5 (68.0-78.0) | <0.001 |
| **Sex** |  |  |  | 0.004 |
| **Female** | 129 (64.2%) | 117 (68.4%) | 12 (40.0%) |  |
| **Male** | 72 (35.8%) | 54 (31.6%) | 18 (60.0%) |  |
| **Education (years)** | 14 (12-17) | 14 (12-16) | 16 (14-18) | <0.001 |
| **Race** |  |  |  | <0.001 |
| **Asian** | 3 (1.5%) | 3 (1.8%) | 0 (0%) |  |
| **B/AA** | 105 (52.2%) | 99 (57.9%) | 6 (20.0%) |  |
| **NHW** | 93 (46.3%) | 69 (40.4%) | 24 (80.0%) |  |
| **MOCA** | 25 (23-27) | 25 (23-27) | 26 (23-27) | 0.427 |
| **APOE ε4 carrier** |  |  |  | 0.131 |
| **Yes** | 61 (30.4%) | 48 (28.1%) | 13 (43.3%) |  |
| **No** | 139 (69.2%) | 122 (71.4%) | 17 (56.7%) |  |
| **Missing** | 1 (0.5%) | 1 (0.6%) | 0 (0%) |  |
| **PiB SUVR** | 1.10 (1.05-1.17) | 1.09 (1.05-1.14) | 1.66 (1.51-1.91) | <0.001 |
| **N Status** |  |  |  | 0.216 |
| **Negative** | 129 (64.2%) | 113 (66.1%) | 16 (53.3%) |  |
| **Positive** | 72 (35.8%) | 58 (33.9%) | 14 (46.7%) |  |
| **Cortical thickness** | 2.74 (2.66-2.79) | 2.74 (2.67-2.80) | 2.71 (2.58-2.76) | 0.018 |

**c. Cohort: HeartSCORE**
|  | **HeartSCORE COHORT** |  |  |  |
| --- | --- | --- | --- | --- |
|  | **Total** | **Aβ PET-** | **Aβ PET+** | **p-value** |
| **N (%)** | 147 | 121 (82.3%) | 26 (17.7%) |  |
| **Age (years)** | 73.0 (70.0-77.0) | 73.0 (70.0-76.0) | 75.0 (70.3-79.0) | 0.206 |
| **Sex** |  |  |  | 0.497 |
| **Female** | 99 (67.4%) | 83 (68.6%) | 16 (61.5%) |  |
| **Male** | 48 (32.7%) | 38 (31.4%) | 10 (38.5%) |  |
| **Education (years)** | 16.0 (13.0-18.0) | 16.0 (14.0-18.0) | 16 (12.0-18.0) | 0.642 |
| **Race** |  |  |  | 0.340 |
| **NHW** | 105 (71.4%) | 84 (69.4%) | 21 (80.8%) |  |
| **B/AA** | 42 (28.6%) | 37 (30.6%) | 5 (19.2%) |  |
| **MMSE** |  |  |  | 0.585 |
| **≥24** | 134 (91.1%) | 109 (90.1%) | 25 (100.0%) |  |
| **19-23** | 5 (3.4%) | 5 (4.1%) | 0 (0%) |  |
| **Missing** | 4 (2.7%) | 4 (3.3%) | 0 (0%) |  |
| **CDR global score** |  |  |  | 0.071 |
| **CDR=0** | 108 (73.5%) | 93 (76.9%) | 15 (57.7%) |  |
| **CDR=0.5** | 23 (15.7%) | 17 (14.1%) | 6 (23.1%) |  |
| **CDR=1.0** | 1 (0.7%) | 1 (0.8%) | 0 (0%) |  |
| **Missing** | 15 (10.2%) | 10 (8.3%) | 5 (19.2%) |  |
| **APOE ε4 carrier** |  |  |  | 0.002 |
| **Yes** | 40 (27.2%) | 26 (21.5%) | 14 (53.8%) |  |
| **No** | 107 (72.3%) | 95 (78.5%) | 12 (46.2%) |  |
| **PiB SUVR** | 1.17 (1.13-1.24) | 1.15 (1.12-1.18) | 1.60 (1.51-1.78) | <0.001 |
| **Tau PET** |  |  |  | <0.001 |
| **Negative** | 97 (66.0%) | 89 (73.6%) | 8 (30.8%) |  |
| **Positive** | 49 (33.3%) | 32 (26.4%) | 17 (65.4%) |  |
| **Missing** | 1 (0.7%) | 0 (0%) | 1 (3.9%) |  |
| **Tau PET SUVR** | 1.15 (1.11-1.21) | 1.4 (1.10-1.19) | 1.23 (1.16-1.27) | <0.001 |

Abbreviations: NHW, non-Hispanic White; B/AA, Black/African American. Numerical variables are reported as median (interquartile range [IQR]), and categorical variables as count (percentage). P values were calculated using the Wilcoxon rank-sum test for numerical variables and Fisher's exact test for categorical variables, with missing values excluded. Tau pathology (T) was assessed using the PET tracer [ $^{18}\text{F}$ ] AV-1451, with a standardized uptake value ratio (SUVR) cutoff of 1.18 to define tau positivity ( $\geq 1.18$ as positive).

The CoBrA cohort had a median (IQR) age of 62 (57-71) years, comprising 129 (64.2%) female participants, 61 (30.4%) *APOE* ε4 allele carriers, 30 (14.9%) A+ participants, and 72 (35.8%) N+ participants (Table 1b). Among them, 105 (52.2%) identified themselves as Black/African American (B/AA), 3 (1.5%) as Asian, and the rest as NHW. The A+ group was older (p<0.001), had fewer female participants (p=0.004), and had a higher likelihood of being NHW (p<0.001) than the A-group, but the two groups had no significant difference in terms of *APOE* ε4 carriership or both dichotomized and numerical N status (Table 1b).

The HeartSCORE cohort had a median (IQR) age of 73 (70-77) years, 99 (67.4%) were females, 40 (27.2%) were *APOE* ε4 carriers. Additionally, 26 (17.7%) were A+, 49 (33.3%) were T+, 39 (26.5%) were N+, and 108 (73.5%) were dementia-free, with a CDR global score of 0 (Table 1c). Among them, 105 (71.4%) were self-identified as NHW and 42 (28.6%) were B/AA. There was no significant difference in terms of age, sex, years of education, or race between the A+ and A-groups, but there was a difference in *APOE* ε4 carrier status (p=0.002), with a higher proportion in the A+ group. Similar to the MYHAT-NI cohort, A+ participants had a higher likelihood of being T+ but not N+.

We also classified the participants based on their tau pathology, and the results are presented in Supplementary Table S1. Among all variables examined, the T+ and T-participants showed significant differences only in their Aβ PET status (both dichotomous and continuous) and Tau PET SUVR.

Significant differences in plasma p-tau217+ levels were observed across the three cohorts, with a p value of < 0.001 in the Kruskal-Wallis test (Figure S1). Post hoc analysis revealed that the MYHAT-NI cohort differed most notably from both the CoBrA and HeartSCORE cohorts.

MYHAT-NI exhibited higher overall p-tau217+ levels, with a median (IQR) of 0.052 (0.037– 0.074) pg/ml, compared with 0.035 (0.026–0.050) pg/ml in CoBrA and 0.038 (0.027–0.055) pg/ml in HeartSCORE. No significant difference was found between the CoBrA and HeartSCORE cohorts. The significant difference across cohorts remained after adjusting for common demographic/genotypic factors, including age, sex, race, education, and *APOE* ε*4* carrier status, with an adjusted N-way ANOVA p-value of 0.012.

### 3.2 Association with amyloid pathology

#### Association with Aβ PET positivity: individual cohorts

We first evaluated the association of plasma p-tau217+ with dichotomized Aβ PET status, separately for each cohort (Figure 1). Significant differences in p-tau217+ levels were observed when comparing A+ to A-groups across all three cohorts, with Wilcoxon rank-sum p-values <0.001 in all three cohorts (Figure 1A). Compared to the A-participants, p-tau217+ levels were 1.80-fold, 2.11-fold, and 1.81-fold higher in the A+ participants for MYHAT-NI, CoBrA, and HeartSCORE, respectively.

**Figure 1.**
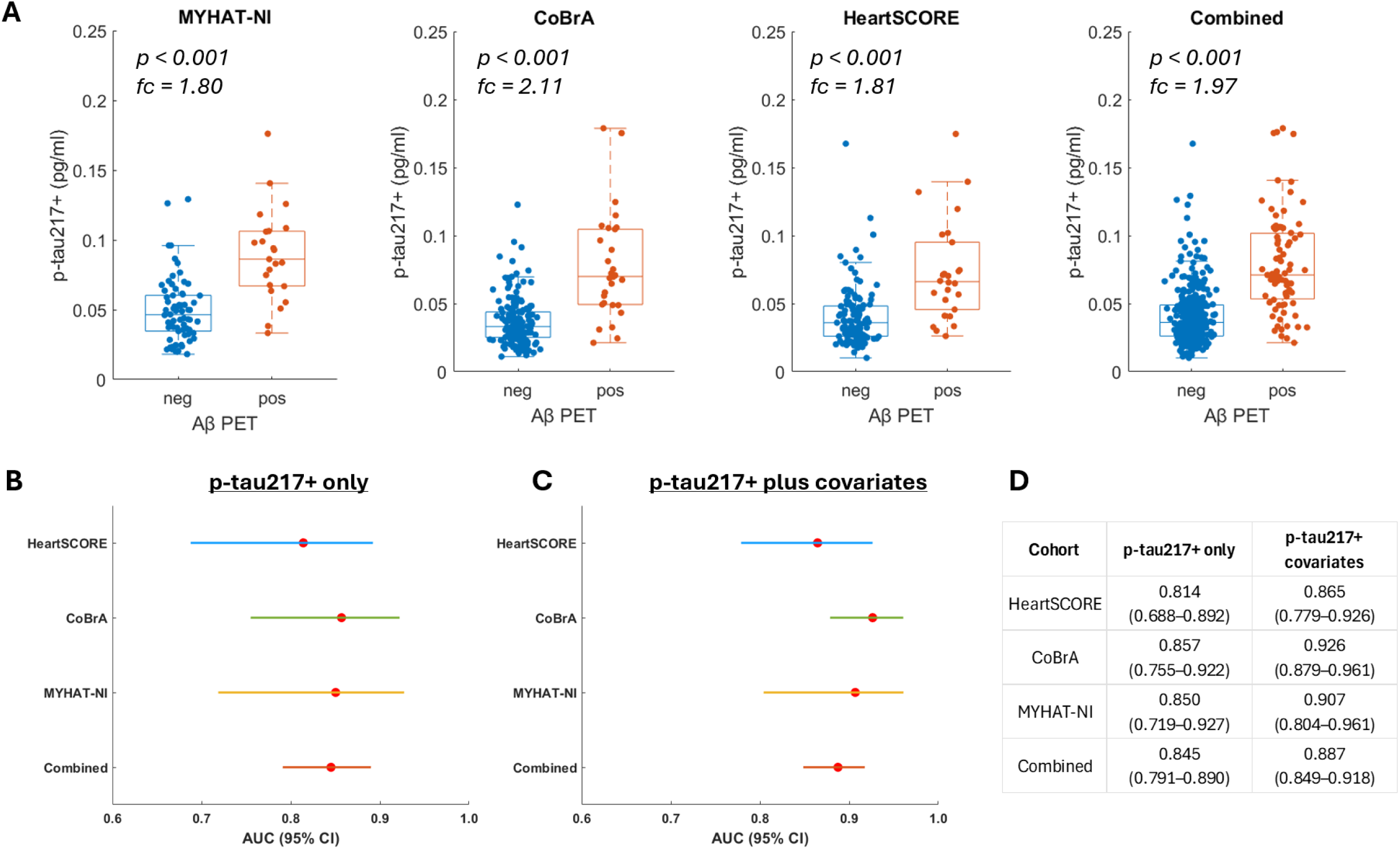
Association of plasma p-tau217+ with Aβ-PET status. Panel A illustrates the boxplot distribution of p-tau217+ levels by Aβ-PET status, with overlaid data points. The central box represents the interquartile range (IQR), the line inside the box indicates the median, and the whiskers extend to the most extreme data points within 1.5 times the IQR from the quartiles. P values were determined using the Wilcoxon rank-sum test, and fold change (fc) represents the ratio of mean p-tau217+ levels in the A+ group relative to the A− group. Panels (B–D) present results from receiver operating characteristic (ROC) analyses evaluating the ability of p-tau217+ to discriminate Aβ-PET positivity across the CoBrA, HeartSCORE, MYHAT-NI, and combined cohorts. Panel B shows a forest plot of area under the curve (AUC) values for p-tau217+ only models. Panel C displays a forest plot of AUC values for models including p-tau217+ along with covariates (age, sex, race, and APOE ε4 carrier status). Panel D presents a table summarizing AUC values and their corresponding 95% confidence intervals (CIs).

We utilized ROC analysis to evaluate the predictive performance of p-tau217 in distinguishing A+ participants from A-individuals. Strong predictive performance was observed in all three cohorts, with AUCs of 0.857 (95% confidence interval [CI]: 0.755 – 0.922), 0.850 (95% CI: 0.719 – 0.927), and 0.814 (95% CI: 0.688 – 0.892) for CoBrA, MYHAT-NI, and HeartSCORE, respectively, based on the p-tau217+ alone (Figure 1B). Inclusion of covariates, including sex, age, race, and *APOE* ε4 carrier status, improved the AUCs of CoBrA to 0.926 (95% CI: 0.879 – 0.961), MYHAT-NI to 0.907 (95% CI: 0.804 – 0.961), and HeartSCORE to 0.865 (95% CI: 0.779 – 0.926) (Figure 1C).

#### Cutoff value estimation

Table 2 shows the classification performance based on cutoff values determined using the Youden index. When using p-tau217+ alone, the optimal thresholds ranged from 0.049 to 0.064 pg/mL, yielding high negative predictive values (NPV: 0.929–0.966) but relatively modest positive predictive values (PPV: 0.463–0.559), with overall accuracy ranging from 0.796 to 0.837. Incorporating covariates generally improved classification performance, except in the HeartSCORE cohort, where accuracy declined from 0.837 to 0.653 after adjusting for covariates.

**Table 2:**
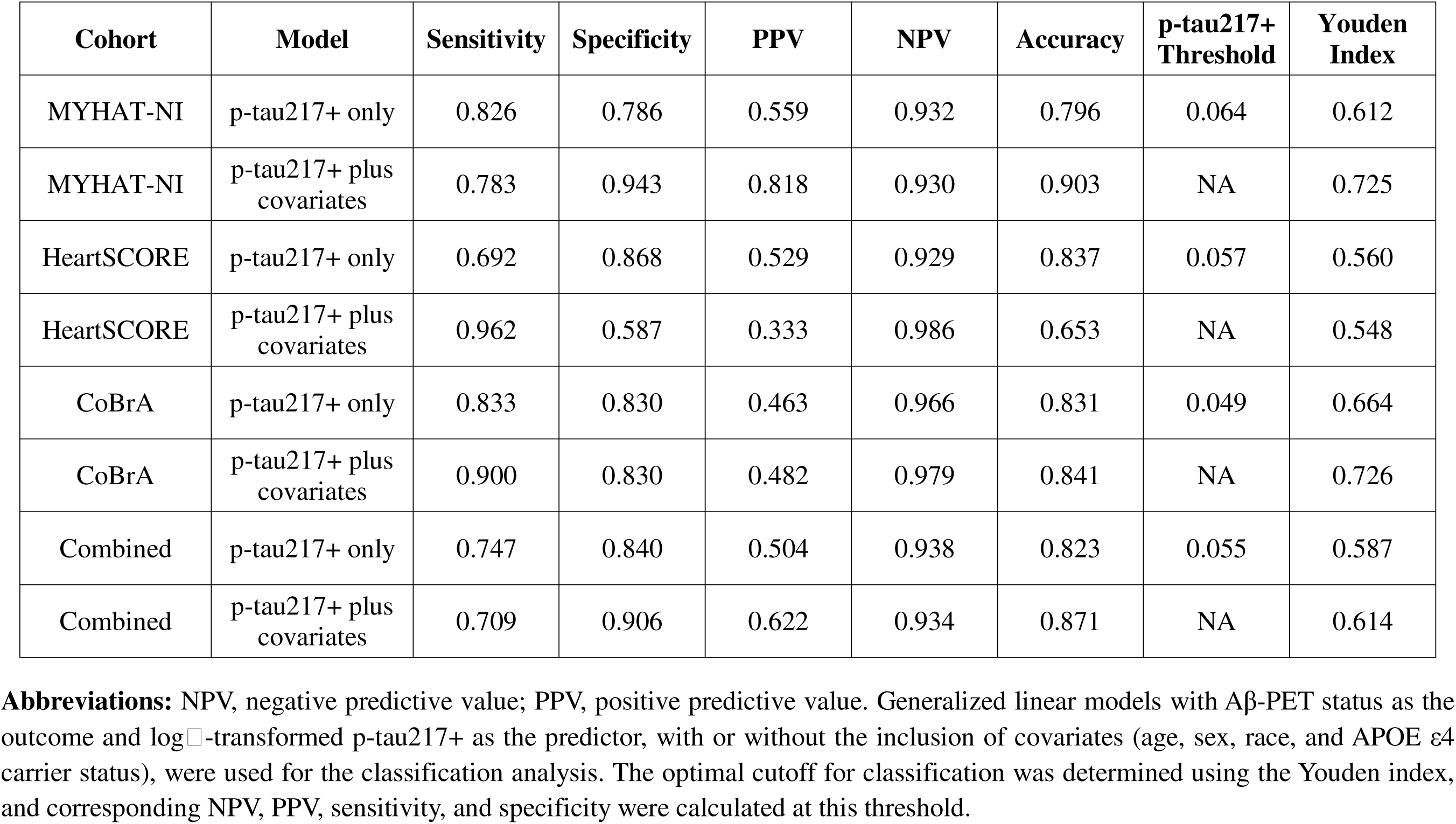
Classification Performance of p-tau217+ Assay in Predicting Amyloid PET Positivity.

| Cohort | Model | Sensitivity | Specificity | PPV | NPV | Accuracy | p-tau217+ Threshold | Youden Index |
| --- | --- | --- | --- | --- | --- | --- | --- | --- |
| MYHAT-NI | p-tau217+ only | 0.826 | 0.786 | 0.559 | 0.932 | 0.796 | 0.064 | 0.612 |
| MYHAT-NI | p-tau217+ plus covariates | 0.783 | 0.943 | 0.818 | 0.930 | 0.903 | NA | 0.725 |
| HeartSCORE | p-tau217+ only | 0.692 | 0.868 | 0.529 | 0.929 | 0.837 | 0.057 | 0.560 |
| HeartSCORE | p-tau217+ plus covariates | 0.962 | 0.587 | 0.333 | 0.986 | 0.653 | NA | 0.548 |
| CoBrA | p-tau217+ only | 0.833 | 0.830 | 0.463 | 0.966 | 0.831 | 0.049 | 0.664 |
| CoBrA | p-tau217+ plus covariates | 0.900 | 0.830 | 0.482 | 0.979 | 0.841 | NA | 0.726 |
| Combined | p-tau217+ only | 0.747 | 0.840 | 0.504 | 0.938 | 0.823 | 0.055 | 0.587 |
| Combined | p-tau217+ plus covariates | 0.709 | 0.906 | 0.622 | 0.934 | 0.871 | NA | 0.614 |

**Abbreviations:** NPV, negative predictive value; PPV, positive predictive value. Generalized linear models with Aβ-PET status as the outcome and log □ -transformed p-tau217+ as the predictor, with or without the inclusion of covariates (age, sex, race, and APOE ε4 carrier status), were used for the classification analysis. The optimal cutoff for classification was determined using the Youden index, and corresponding NPV, PPV, sensitivity, and specificity were calculated at this threshold.

#### Correlation with Aβ burden measured by PiB SUVR

As depicted in Figure 2, plasma p-tau217+ levels showed a strong positive correlation with PiB SUVR across all three cohorts, with Spearman correlation coefficients (ρ) of 0.336 (p=0.001), 0.384 (p<0.001), and 0.397 (p<0.001) for MYHAT-NI, CoBrA, and HeartSCORE, respectively. Overall, a stronger correlation was observed among A+ participants, with ρ values of 0.426 (0.044), 0.333 (p=0.073), and 0.642 (p<0.001) for MYHAT-NI, CoBrA, and HeartSCORE, respectively. The corresponding ρ values among A-participants were -0.136 (p=0.262), 0.162 (p=0.035), and 0.164 (p=0.072).

**Figure 2:**
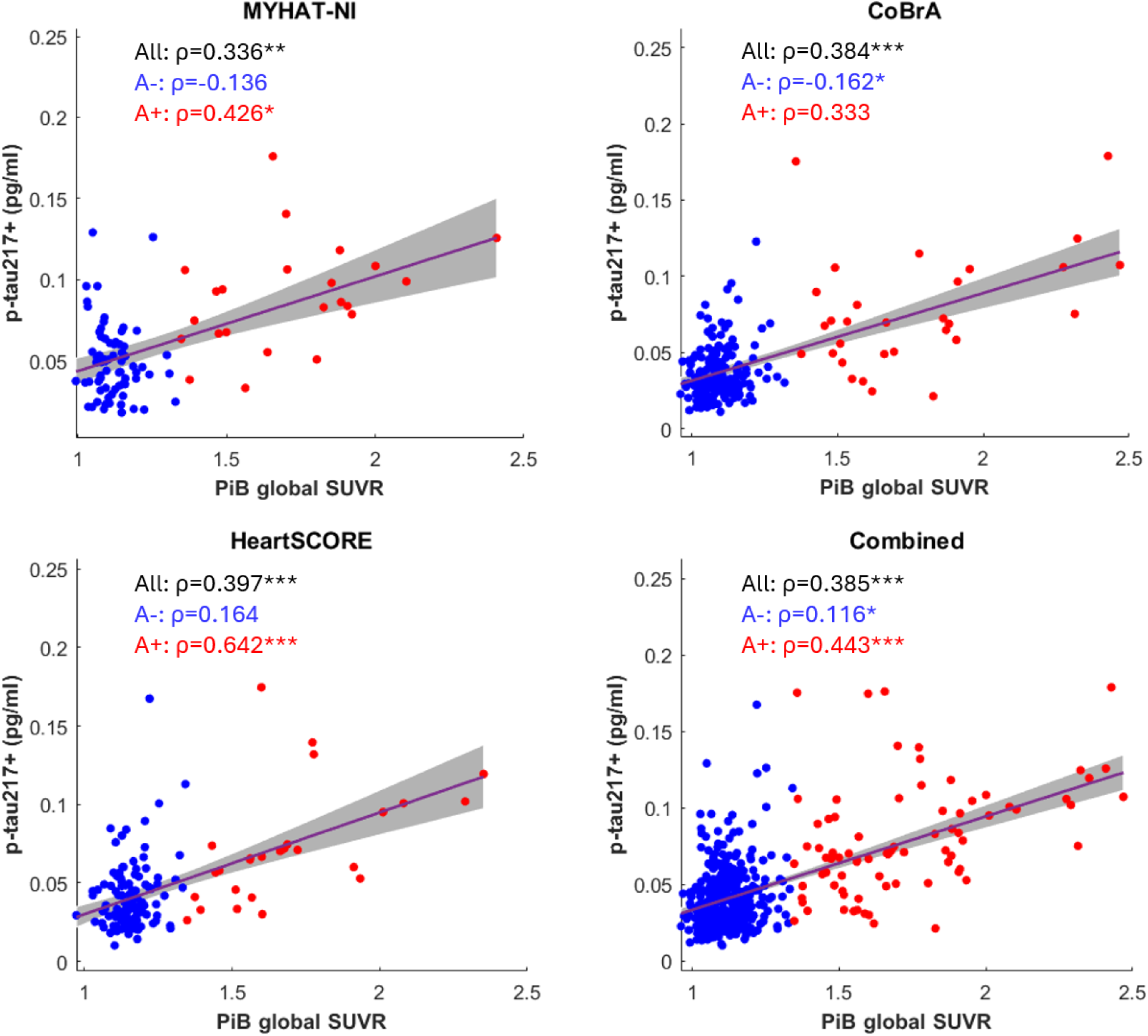
Scatterplots depict the correlation between plasma p-tau217+ levels and amyloid burden measured by PiB SUVR. Spearman correlation was used to assess associations. Blue dots represent A− individuals, and red dots represent A+ individuals. The red line indicates the linear regression across the entire cohort, with the gray shaded area representing the 95% confidence interval. Spearman’s rho (ρ) values are shown for A-, A+, and the entire cohort. Asterisks indicate significance levels: *, p < 0.05; **, p < 0.01; ***, p < 0.001.

#### Association in the combined cohort

When combining all three cohorts, the overall fold change in p-tau217+ levels between A+ and A− groups was 1.97. The AUC for distinguishing A+ from A− individuals was 0.845 (95% CI: 0.791–0.890) using p-tau217+ alone and improved to 0.887 (95% CI: 0.849–0.918) when covariates were included. Consistent with findings from individual cohorts, the optimal threshold determined by Youden index yielded a high NPV but less robust PPV. Specifically, the NPV and PPV were 0.938 and 0.504, respectively, using p-tau217+ alone, and 0.934 and 0.622 when covariates were incorporated. The combined cohort gave a threshold of 0.05 pg/ml. Although p-tau217+ levels were elevated in the MYHAT-NI cohort compared to CoBrA and HeartSCORE, there was no significant cohort-specific association with Aβ PET status, as indicated by the non-significant interaction between cohort and A status on p-tau217+ levels (data not shown).

### 3.3 Association with tau pathology

We further analyzed the correlation between plasma p-tau217+ and tau PET status to determine its potential for detecting tau neurofibrillary tangle pathology (Figure 3). A similar fold change difference was observed when comparing T+ vs. T-individuals for MYHAT-NI, HeartSCORE, and the combined cohorts, with values of 1.31 (p=0.032), 1.34 (p=0.007), and 1.33 (p<0.001), respectively. ROC analysis revealed modest diagnostic performance, with AUC values of 0.635 (95% CI: 0.494-0.746) for MYHAT-NI and 0.638 (95% CI: 0.538-0.722) for HeartSCORE, when using plasma p-tau217+ only. Incorporating covariates led to slight improvements, with AUCs increasing to 0.666 (95% CI: 0.533-0.767) and 0.654 (95% CI: 0.558-0.749), respectively. When the two cohorts were combined, the AUC was 0.633 (95% CI: 0.571-0.681) using plasma p-tau217+ alone, and 0.646 (95% CI: 0.592-0.697) after adjusting for covariates.

**Figure 3:**
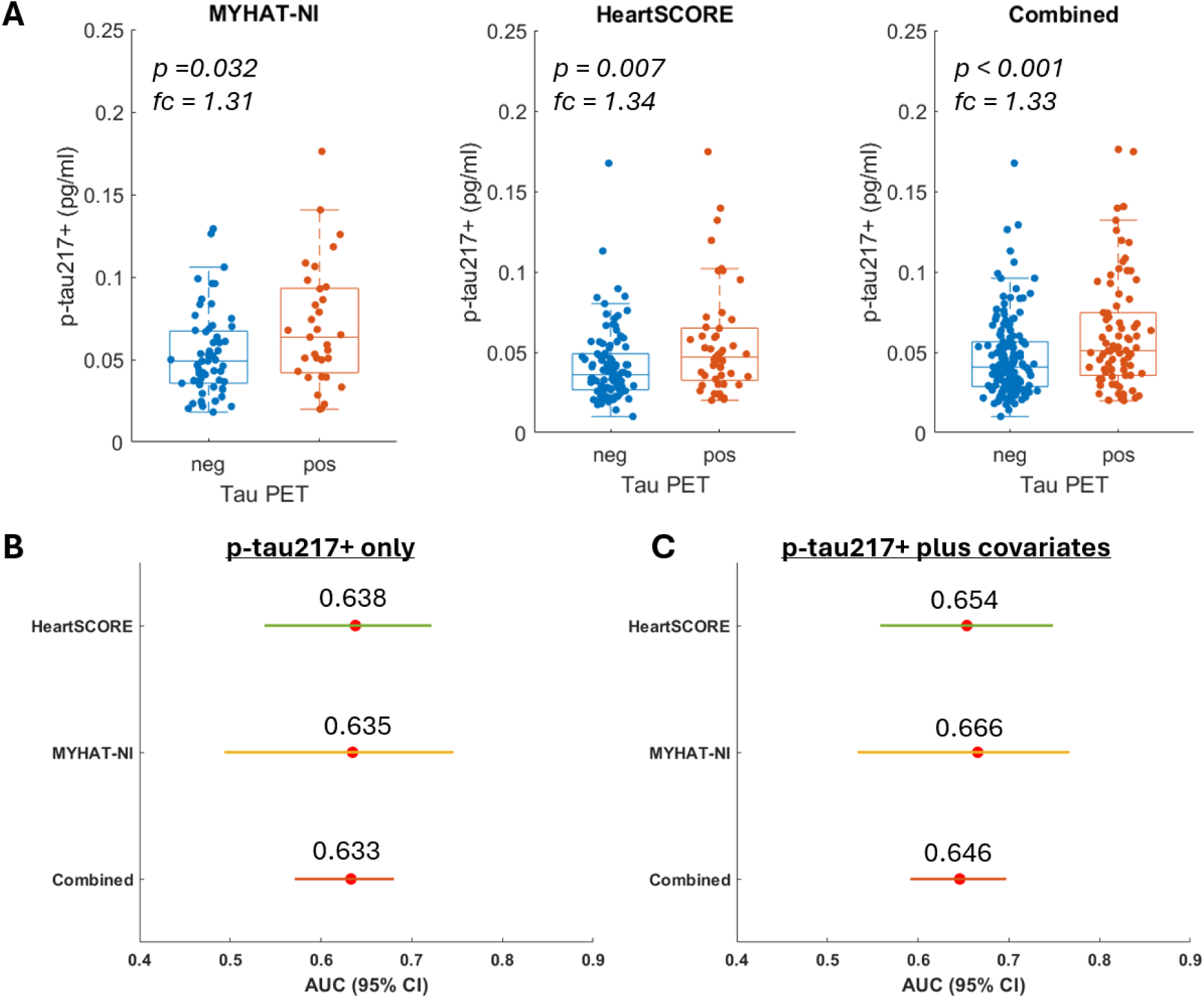
Association of plasma p-tau217+ with Tau PET status. Panel A illustrates the boxplot distribution of p-tau217+ levels by Tau PET status, with overlaid data points. P values were determined using the Wilcoxon rank-sum test, and fold change (fc) represents the ratio of mean p-tau217+ levels in the T+ group relative to the T− group. Panels (B–C) present results from receiver operating characteristic (ROC) analyses evaluating the ability of p-tau217+ to discriminate Tau PET positivity across the HeartSCORE, MYHAT-NI, and combined cohorts. Panel B shows a forest plot of AUC values for p-tau217+ only models. Panel C displays a forest plot of AUC values for models including p-tau217+ along with covariates (age, sex, race, and *APOE* ε4 carrier status).

We then evaluated the strength of the association between plasma p-tau217+ levels and tau tangle load determined by tau PET SUVR (Figure 4). P-tau217+ levels showed a moderate correlation with Tau-PET SUVR, with ρ values of 0.195 (p=0.061), 0.336 (p<0.001), and 0.265 (p<0.001) for MYHAT-NI, HeartSCORE, and combined cohort, respectively. Similar to the association with Aβ burden, a stronger correlation was observed among T+ participants, with ρ values of 0.354 (p=0.044), 0.466 (p<0.001), and 0.384 (p<0.001) for MYHAT-NI, HeartSCORE, and combined cohort, respectively. The corresponding ρ values among T-participants were - 0.080 (p=0.544), 0.227 (p=0.025) and 0.095 (p=0.239).

**Figure 4:**
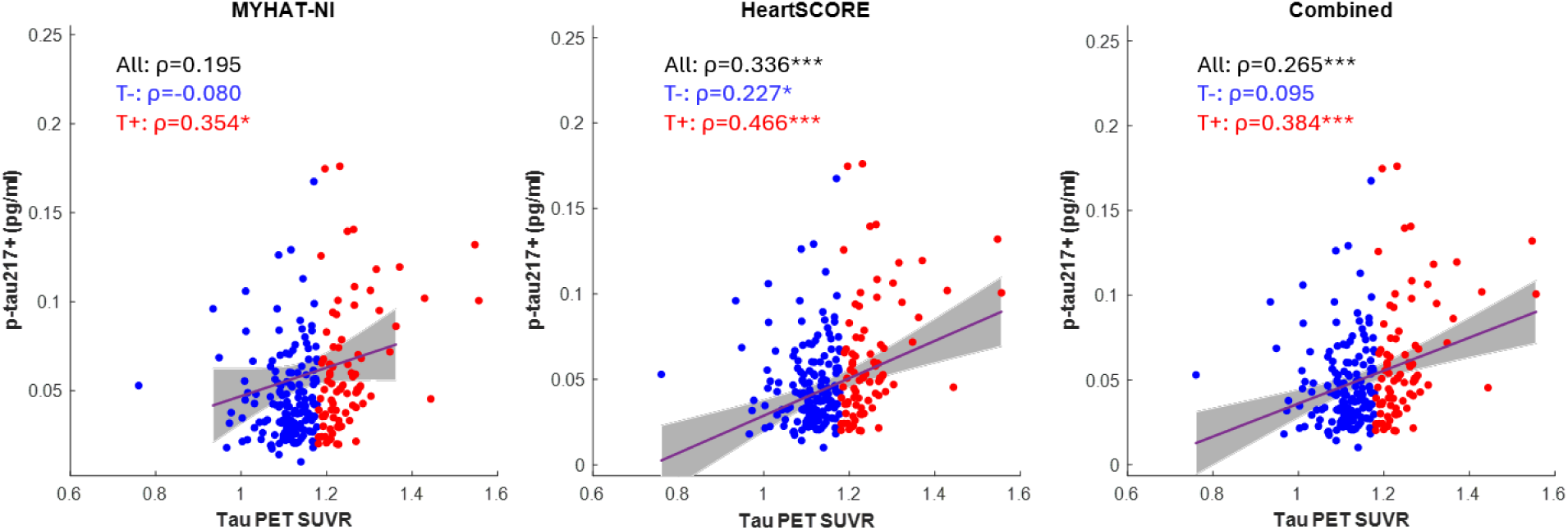
Scatterplots depict the correlation between plasma p-tau217+ levels and tau tangle load measured by tau PET SUVR. Spearman correlation was used to assess associations. Blue dots represent T− individuals, and red dots represent T+ individuals. The red line indicates the linear regression across the entire cohort, with the gray shaded area representing the 95% confidence interval. Spearman’s rho (ρ) values are shown for T-, T+, and the entire cohort. Asterisks indicate significance levels: *, p < 0.05; **, p < 0.01; ***, p < 0.001.

### 3.4 Plasma p-tau217+ levels by A/T classification

To gain a better understanding of how plasma p-tau217+ varies across different stages of AD pathology, we compared its profiles across four groups stratified by A and T status. As shown in Figure 5, median p-tau217+ levels increased stepwise across the A-T-, A-T+, A+T-, and A+T+ groups in both individual and combined cohorts. However, in the individual cohorts, significant differences were observed only between A+T+ and A-T- or A-T+ (all p < 0.001). In the combined cohort, the A+T- also differed significantly from the A-T- (p = 0.005) and A-T+ (p = 0.019) groups. No significant differences were found between T- and T+ groups when A status was held constant (i.e., A-T- vs. A-T+ or A+T- vs. A+T+), suggesting that A status primarily drives the observed changes in p-tau217+ levels. Consistent with this, T status was not significantly associated with p-tau217+ levels after adjusting for A status in either cohort (data not shown).

**Figure 5:**
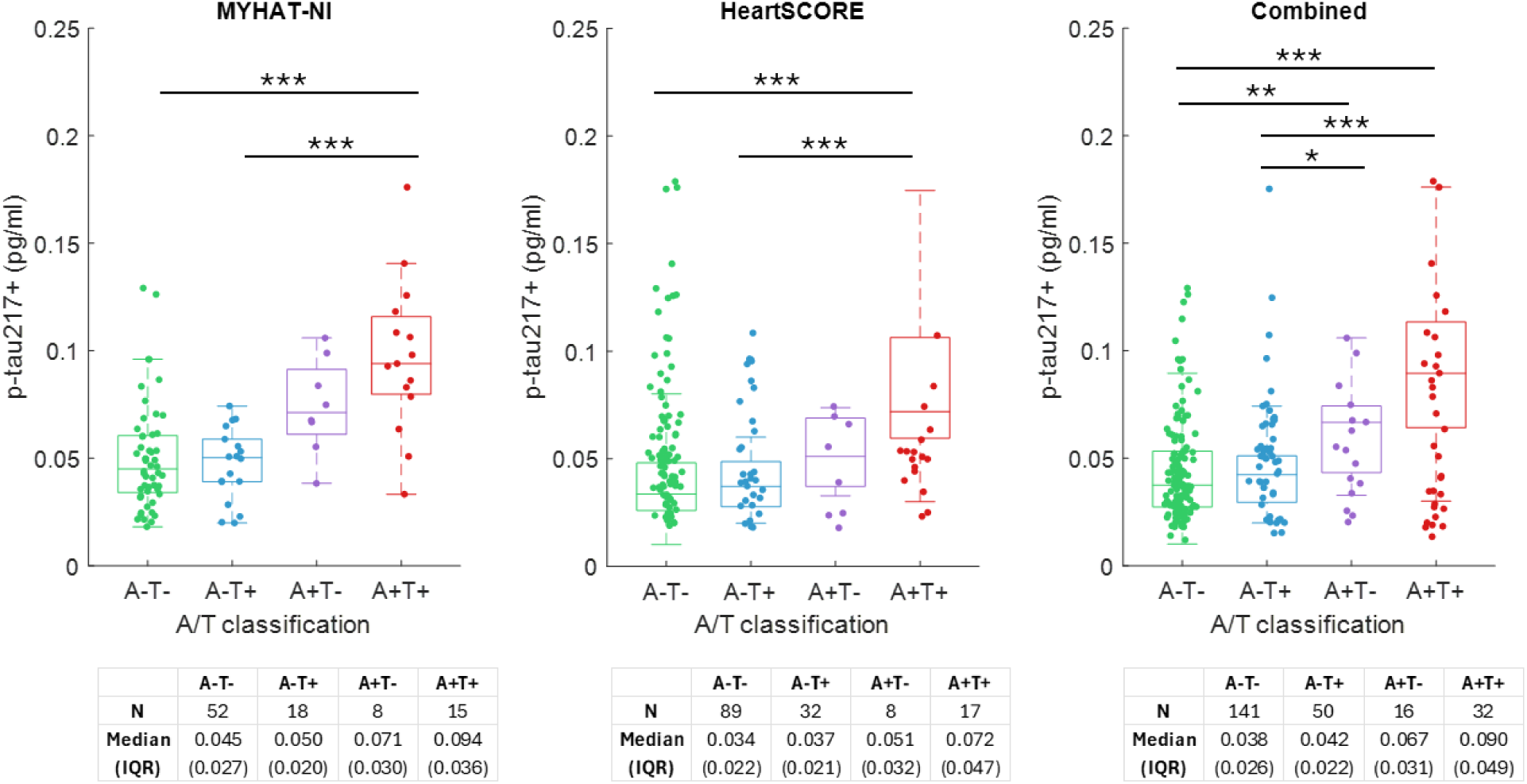
Plasma p-tau217+ profiles by A/T classification. P values were calculated using post hoc analysis following the Kruskal–Wallis test, with Bonferroni correction for multiple comparisons. Asterisks indicate significance levels: *, p < 0.05; **, p < 0.01; ***, p < 0.001.

### 3.5 Association of p-tau217+ with cognitive impairment

We evaluated the association of p-tau217+ with cognitive impairment using different assessment tools across three cohorts: the CDR global score in MYHAT-NI (CDR 0 = normal; CDR 0.5 = impaired), the MoCA in CoBrA (≥26 = normal; <26 = impaired), and the MMSE in HeartSCORE (≥24 = normal; <24 = impaired). As shown in Figure S2, among the three cohorts, only the MYHAT-NI cohort demonstrated a marginally significant elevation in p-tau217+ levels in the impaired group, with a fold change of 1.38 (p = 0.086). In contrast, both CoBrA and HeartSCORE showed lower p-tau217+ levels in the impaired groups, with fold changes of 0.581 (p = 0.996) and 0.838 (p = 0.527), respectively; however, these differences were not statistically significant.

### 3.6 Associations of plasma p-tau217+ levels with demographic and genetic risk factors

Age, sex, race, education, and *APOE* ε*4* genotype are commonly recognized as important confounding factors in AD research. Table S2 illustrates the relationship between p-tau217+ levels and these factors in our cohorts. In univariate analyses, we observed a significant positive correlation between age and p-tau217+ levels in both the MYHAT-NI and CoBrA cohorts, with marginal significance in the HeartSCORE cohort. *APOE* ε4 carrier status was associated with higher p-tau217+ levels in MYHAT-NI and HeartSCORE (p = 0.028 and 0.014, respectively), but not in CoBrA (p = 0.516). In the CoBrA cohort, race, sex, and education also showed significant univariate associations with p-tau217+ levels. However, in multivariable models that included all these factors, only age remained significantly associated with p-tau217+ levels (p<0.001), while sex showed marginal significance (p=0.051).

## 4 DISCUSSION

The transformative utility of blood-based biomarkers in the field of AD and related dementias continues to become increasingly realistic due to the introduction of various tests that have high accuracies, facilitating enhanced patient choices and diagnostic market competition for predicting A-T-N status. Among these, the p-tau tests–specifically p-tau217, p-tau231, and p-tau181–have garnered the most attention. Notably, p-tau217 has demonstrated the highest reliability and potential for diagnosing AD. Companies such as Lilly, ALZpath, C2N, Quanterix, Alamar, Roche, Janssen, Beckman Coulter, and Fujirebio have developed p-tau217 assay platforms that show promising results for clinical applications. The Janssen plasma p-tau217+ assay used in this study has yielded results comparable to the ALZpath p-tau217 in a translational study cohort, showing utility for monitoring and detecting AD pathology.^15^ In another study comparing multiple plasma p-tau217 assays, the Janssen p-tau217+ assay demonstrated a strong capability to identify abnormal Aβ and tau PET status.^13^ Janssen has also transferred non-exclusive commercialization rights for its p-tau217+ assay to Quanterix, the same company which manages the ALZpath assay. This assay is now commercially available as a Lab Developed Test through Quanterix as the LucentAD p217 assay.^23^

This study evaluated the Janssen plasma p-tau217+ assay in individuals mostly in the preclinical phase of AD pathology using three independent community-based cohorts. We examined the assay’s diagnostic capability for detecting AD and its association with both Aβ and tau PET. Plasma p-tau217+ differentiated Aβ PET-negative from Aβ PET-positive participants, as well as when classified according to combined Aβ and tau PET status. When the amyloid and tau statuses were stratified together, the results showed that the A status was the main driver of the changes seen in the p-tau217+ assay as the levels increased from A-T- to A-T+ to A+T-, and finally A+T+. This pattern was consistent across cohorts. The differences between A+ and A-participants were consistently observed across the cohorts and the AUCs for the Aβ PET were comparable, indicating reliability for classification purpose. This underscores a strong relationship between p-tau217+ and Aβ burden examined on the Aβ PET scan.

In contrast, plasma p-tau217+ performed moderately in correlating with tau PET results compared to Aβ PET results. While plasma p-tau217+ showed significant difference between T+ and T-participants, it only achieved an average AUC of about 65%. In addition, T status no longer showed a significant association with p-tau217+ levels after adjusting for A status. This finding aligns with previous reports indicating that further understanding of site-specific patterns of tau phosphorylation is necessary before the p-tau217+ assay can be considered as a potential surrogate for tau PET.^11,24^ Ongoing effort has led to the discovery of other assays such as p-tau serine-262 and serine-356 which correlate more strongly with tau PET.^25^ In contrast to our findings, Warmenhoven et al.,^23^ reported higher AUCs and accuracies for the Janssen plasma p-tau217+ assay in identifying individuals with abnormal tau PET status. The discrepancies in our outcomes may arise from the fact that our study comprised community-setting cohorts, while the other study focused on participants from a memory clinic with a higher prevalence of AD pathology. While the Warmenhoven study was composed of 48% cognitively impaired participants, our cohorts were composed of less than 10% combined. Additionally, there were only a few participants in the present study cohorts who were tau PET positive. Among those who are, many have low SUVR, which makes it challenging to establish a meaningful correlation between p-tau217+ and Tau PET.

A key takeaway from this study is the high specificity and sensitivity (above 80%) of the Janssen plasma p-tau217+ assay across all cohorts. This is particularly noteworthy given that the vast majority of the cohorts consisted of cognitive normal older adults. In terms of the performance values, there was high NPV which makes p-tau217+ a good tool for the initial screening of high-risk individuals or clinical trial participants. However, the low PPV suggests that an orthogonal test is needed to confirm the presence of AD pathology among those with high p-tau217+. The low PPV could also be attributed to the fact that many participants in the cohorts are cognitively unimpaired. We adopted the optimal cutpoint strategy from the Alzheimer’s Association workgroup revised criteria for diagnosing and staging of AD,^9^ establishing an average threshold of 0.05 pg/ml across cohorts and when the cohorts were combined. To our knowledge, this is the first study to assess the cutoff in the Janssen plasma p-tau217+ assay.

Regarding demographics, the assay effectively highlighted the influence of demographics factors on biomarker testing. When adjusted for covariates such as *APOE* ε4 genotype, education, sex and age, the p-tau217+ assay showed resilient associations within the cohort. This further emphasizes that these covariates have a minimal impact on p-tau217 assay compared to other assays, like p-tau181 which are more influenced by such factors.^12^

Many studies on plasma p-tau217+ have been limited by a lack of diversity, often overlooking its performance in real-world settings. However, this study addresses this gap by examining various ethnicities, races, and socio-economic backgrounds. For instance, all cohorts analyzed differences between NHW and B/AA, while the CoBrA cohort even included Asian participants despite being a very small proportion. In conclusion, this study provides strong evidence that the Janssen plasma p-tau217+ assay is a reliable biomarker for prognostic purposes, even in community settings.

This study has a key limitation: the lack of long-term follow-up data to assess the relationship between the p-tau217+ assay and the progression of AD. The current analysis is based solely on baseline assessment data, highlighting the critical need for follow-up information. Without follow-up data, it is challenging to evaluate the long-term effectiveness and reliability of the p-tau217+ assay as a biomarker for AD pathology. Future research should include follow-up assessments to deepen our understanding and validate the role of the p-tau217+ assay in monitoring AD progression.

While the Janssen plasma p-tau217+ assay correlates with both Aβ and tau pathology, our results show that it is particularly effective in identifying individuals with abnormal Aβ plaques, as indicated by its strong correlation with Aβ PET imaging findings. This study suggests that p-tau217+ could be a valuable biomarker for the early detection of AD pathophysiology with high sensitivity, specificity, and predictive values. Together, this may enable earlier detection, and access to interventions – both pharmacological and non-pharmacological – potentially slowing disease progression.

## Data Availability

All data produced in the present study are available upon reasonable request to the authors

## ACKNOWLEDGMENTS

We thank all study participants, their families and caretakers in this cohort studies. The MYHAT, CoBRA and HeartSCORE studies were funded by NIH grants R37AG023651, R01AG072641, R01AG052521, and R01HL089292. This study used biomarker testing infrastructure established with support from NIH/NIA R01AG083874. TKK and the Karikari Laboratory were further supported by NIH/NIA (U24AG082930, P30AG066468, RF1AG077474, R01AG083156, R01AG025516, R01AG073267, R01AG075336, P01AG025204), NIH/NINDS (U01NS131740, U01NS141777), NIH/NIMH (R01MH108509), Aging Mind Foundation (DAF2255207), the Department of Defense (HT94252320064), the Anbridge Charitable Fund, and a professorial endowment from the Department of Psychiatry, University of Pittsburgh. The content of this article is solely the responsibility of the authors and does not necessarily represent the official views of the funders.

## CONSENT STATEMENT

All the studies were approved by the Institutional Review Board at the University of Pittsburgh. All subjects provided written informed consent.

