## Supplementary data for "Cross-cohort validation and cutpoint estimation of the Janssen plasma p-tau217+ assay in predominantly cognitively normal community cohorts"

**Table S1: Participant characteristics in the MYHAT-NI and HeartSCORE cohorts according to Tau Positivity**

a. **Cohort MYHAT-NI**

|  | **MYHAT-NI COHORT** | | | |
| --- | --- | --- | --- | --- |
|  | **Total** | **Tau PET-** | **Tau PET+** | **p-value** |
| **N (%)** | 93 | 60 (64.5%) | 33 (35.5%) |  |
| **Age (years)** | 76.0 (72.0-81.0) | 77.0 (72.5-80.5) | 76.0 (72.0-83.3) | 0.891 |
| **Sex**  **Female**  **Male** | 48 (51.6%)  45 (48.4%) | 29 (48.3%)  31 (51.7%) | 19 (57.6%)  14 (42.4%) | 0.516 |
| **Education (years)** | 12.0 (12.0-16.0) | 12.0 (12.0-16.0) | 13 (12.0-14.5) | 0.888 |
| **Race**  **NHW**  **B/AA** | 89 (95.7%)  4 (4.3%) | 57 (95.0%)  3 (5.0%) | 32 (97.0%)  1 (3.0%) | 1.000 |
| **MMSE**  **>=24**  **19-23** | 91 (97.9)  2 (2.2%) | 58 (96.7%)  2 (3.3%) | 33 (100.0%)  0 (0.0%) | 0.537 |
| **CDR**  **CDR=0**  **CDR=0.5** | 85 (91.4%)  8 (8.6%) | 55 (91.7%)  5 (8.3%) | 30 (90.9%)  3 (9.1%) | 1.000 |
| ***APOE* ε4 carrier**  **Yes**  **No** | 13 (14.0%)  80 (86.0%) | 5 (8.3%)  55 (91.7%) | 8 (24.2%)  25 (75.8%) | 0.058 |
| **PiB SUVR** | 1.14 (1.08-1.33) | 1.12 (1.07-1.20) | 1.19 (1.10-1.73) | 0.002 |
| **A Status**  **Negative**  **Positive** | 70 (75.3%)  23 (24.7%) | 52 (86.7%)  8 (13.3%) | 18 (54.5%)  15 (45.5%) | <0.001 |
| **Tau PET SUVR** | 1.16 (1.11-1.21) | 1.12 (1.09-1.15) | 1.22 (1.21-1.26) | <0.001 |

b. **Cohort HeartSCORE**

|  | **HeartSCORE COHORT** | | | |
| --- | --- | --- | --- | --- |
|  | **Total** | **Tau PET -** | **Tau PET +** | **p-value** |
| **N (%)** | 146 | 97 (66.4%) | 49 (33.6%) |  |
| **Age** | 73.0 (70.0-77.0) | 73.0 (70.0-77.0) | 73.0 (70.8-76.0) | 0.899 |
| **Sex**  **Female**  **Male** | 98 (67.1%)  48 (32.9%) | 62 (63.9%)  35 (36.1%) | 36 (73.5%)  13 (26.5%) | 0.269 |
| **Education (years)** | 16.0 (13.0-18.0) | 16.0 (13.8-18.0) | 16.0 (12.0-18.0) | 0.272 |
| **Race**  **NHW**  **B/AA** | 104 (71.2%)    42 (28.8%) | 71 (73.2%)   26 (26.8%) | 33 (67.3%)  16 (32.7%) | 0.562 |
| **MMSE**  **> =24**  **19-23**  **Missing** | 133 (91.1%)  5 (3.4%)  8 (5.5%) | 89 (91.8%)  5 (5.2%)  3 (3.1%) | 44 (89.8%)  0 (0.0%)  5 (10.2%) | 0.177 |
| **CDR Global**  **CDR=0**  **CDR=0.5**  **CDR= 1.0**  **Missing** | 107 (73.3%)  23 (15.8%)  1 (0.7%)  15 (10.3%) | 72 (74.2%)  17 (17.5%)  1 (1.0%)  7 (7.2%) | 35 (71.4%)  6 (12.2%)  0 (0.0%)  8 (16.3%) | 0.115 |
| ***APOE* ε4 carrier**  **Yes**  **No** | 39 (26.7%)  107 (73.3%) | 24 (24.7%)  73 (75.3%) | 15 (30.6%)  34 (69.4%) | 0.553 |
| **PiB SUVR** | 1.16 (1.13 -1.24) | 1.15 (1.11-1.20) | 1.21 (1.16-1.60) | <0.001 |
| **A Status**  **Negative**  **Positive** | 121 (82.9%)  25 (17.1%) | 89 (91.8%)  8 (8.2%) | 32 (65.3%)  17 (34.7%) | <0.001 |
| **Tau PET SUVR** | 1.15 (1.11-1.21) | 1.12 (1.09-1.15) | 1.23 (1.21-1.27) | <0.001 |

Abbreviations: NHW, non-Hispanic White; B/AA, Black/African American. Numerical variables are reported as median (interquartile range [IQR]), and categorical variables as count (percentage). P values were calculated using the Wilcoxon rank-sum test for numerical variables and Fisher’s exact test for categorical variables, with missing values excluded. Tau pathology (T) was assessed using the PET tracer [¹⁸F] AV-1451, with a standardized uptake value ratio (SUVR) cutoff of 1.18 to define tau positivity (≥1.18 as positive).

**Table S2: Association of p-tau217+ levels with demographic and genetic factors**

|  | **MYHAT-NI** | | **CoBrA** | | **HeartSCORE** | |
| --- | --- | --- | --- | --- | --- | --- |
|  | **Estimate** | **p value** | **Estimate** | **p value** | **Estimate** | **p value** |
| **Models with individual factors** | | | | | | |
| **Age** | 0.024 | 0.048 | 0.033 | 0.000 | 0.023 | 0.078 |
| **Race** | 0.123 | 0.746 | -0.329 | 0.001 | -0.051 | 0.705 |
| **Sex** | -0.047 | 0.758 | 0.322 | 0.004 | 0.058 | 0.654 |
| **education** | -0.030 | 0.415 | 0.061 | 0.000 | 0.015 | 0.508 |
| ***APOE* ε4 carriership** | 0.480 | 0.028 | 0.076 | 0.516 | 0.331 | 0.014 |
| **Combined model** | | | | | | |
| **Age** | 0.023 | 0.058 | 0.027 | 0.000 | 0.027 | 0.033 |
| **Race** | -0.009 | 0.981 | -0.111 | 0.298 | -0.045 | 0.741 |
| **Sex** | 0.013 | 0.931 | 0.200 | 0.051 | 0.073 | 0.568 |
| **education** | -0.026 | 0.474 | 0.012 | 0.466 | 0.025 | 0.271 |
| ***APOE* ε4 carriership** | 0.486 | 0.030 | 0.113 | 0.292 | 0.368 | 0.007 |

P values were determined using linear regression models with log2-transformed p-tau217+ levels as the outcome variable and the corresponding factor(s) as predictor(s). For the combined model, all listed factors were included simultaneously in a single model for each cohort.

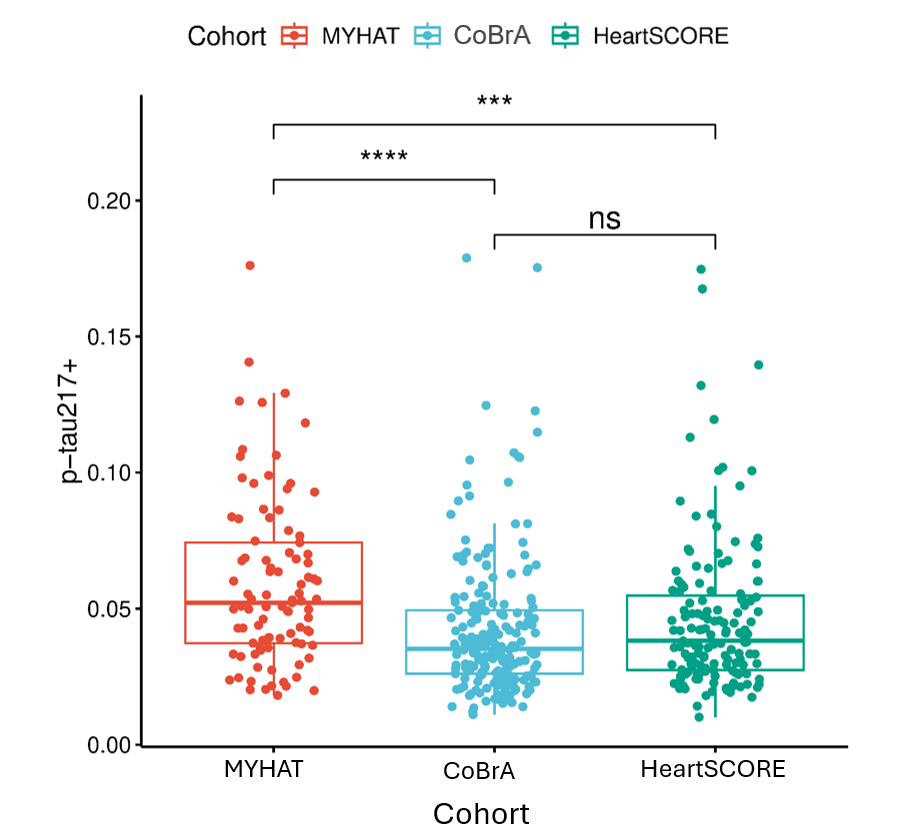

**Figure S1: Boxplot distribution for p-tau217 levels across three cohorts.** Pairwise comparisons were conducted using post hoc analysis following the Kruskal-Wallis test, with Bonferroni correction applied for multiple comparisons. Asterisks denote significance levels: ***, p < 0.001; ****, p < 0.0001.

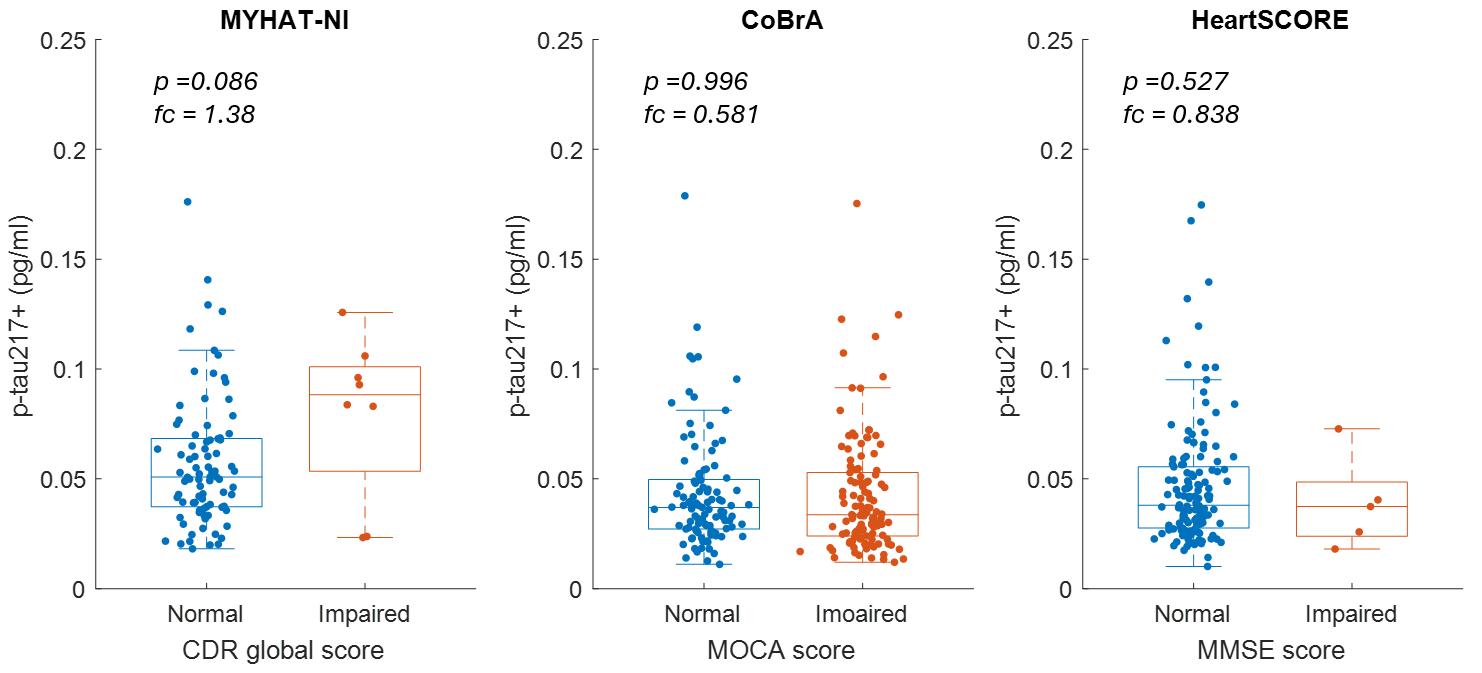

**Figure S2: Association of p-tau217+ with cognitive impairment.** P values were determined using the Wilcoxon rank-sum test, and fold change (fc) represents the ratio of mean p-tau217+ levels in the impaired group relative to the normal group
